# Resting-state EEG distinguishes depression in Parkinson’s disease

**DOI:** 10.1101/2022.02.16.22271060

**Authors:** Arturo I Espinoza, Patrick May, Md Fahim Anjum, Arun Singh, Rachel C Cole, Nicholas Trapp, Soura Dasgupta, Nandakumar S Narayanan

## Abstract

**Introduction:** Depression is a non-motor symptom of Parkinson’s disease (PD). PD-related depression is hard to diagnose and the neurophysiological basis is poorly understood. Depression can markedly affect cortical function, which suggests that scalp electroencephalography (EEG) may be able to distinguish depression in PD.

**Methods:** We recruited 18 PD patients, 18 PD patients with depression, and 12 demographically-similar non-PD patients with clinical depression. All patients were on their usual medications. We collected resting-state EEG in all patients and compared cortical brain signal features between patients with and without depression. We used a machine-learning algorithm that harnesses the entire power spectrum (linear predictive coding of EEG Algorithm for PD: LEAPD), to distinguish between groups.

**Result:** We found differences between PD patients with and without depression in the alpha band (8-13 Hz) globally and in the beta (13-30 Hz) and gamma (30-80 Hz) bands in the central electrodes. From two minutes of resting-state EEG we found that LEAPD-based machine learning could robustly distinguish between PD patients with and without depression with 97% accuracy, and between PD patients with depression and non-PD patients with depression with 100% accuracy. We verified the robustness of our finding by confirming that the classification accuracy declines gracefully as data are truncated.

**Conclusions:** We demonstrated the efficacy of the LEAPD algorithm in identifying PD patients with depression from PD patients without depression and controls with depression. Our data provide insight into cortical mechanisms of depression and could lead to novel neurophysiologically-based biomarkers for non-motor symptoms of PD.

**HIGHLIGHTS:**

- We used EEG to analyze depression in Parkinson’s disease.
- Depressed Parkinson’s patients had distinct spectral EEG features.
- Machine-learning algorithms could accurately distinguish depression in Parkinson’s disease.

## INTRODUCTION

Depression is a prominent non-motor symptom of Parkinson’s disease (PD) [1]. PD-related depression affects ∼20%–40% of PD patients, several times the expected prevalence within this population [2]. Importantly, this aspect of PD is often missed by physicians, contributing to morbidity and decreased quality of life [3–6]. Despite its significance and impact [7], it is unclear which brain circuits contribute to PD-related depression [8]. Determining which brain circuits are involved could lead to the development of new diagnostic tools to identify PD-related depression, as well as targeted treatments such as neuromodulation [9]. A fast and accurate neurophysiologically-based diagnostic tool may also facilitate neuromodulation. In addition, a better understanding of depression in PD may help us illuminate fundamental mechanisms of both diseases. PD and depression involve several overlapping circuits and associated neurotransmitters, including dopamine and serotonin [10]. These projection systems affect cortical physiology [11,12]. Cortical regions can be profoundly dysfunctional in PD [13] and in depression [14].

One technique that is particularly well-suited to capture cortical neurophysiology is electroeencephalography (EEG), which uses scalp electrodes to record activity from the cortex via an array of scalp electrodes. An early EEG study comparing depressed and non-depressed PD patients found widespread differences in alpha bands (8-13 Hz) in posterior and frontal sites [7]. Quantititave EEG (qEEG) studies have found spectral differences that distinguished PD vs depresson [15]. Furthermore, prefrontal cortical regions are responsive to targeted interventions, such as transcranial magnetic stimulation [16]. Here, we tested the hypothesis that spectral features of EEG can distinguish PD patients with depression.

We tested this hypothesis by collecting resting-state scalp EEG in PD patients with and without depression. We compared these data with control patients with depression but without PD. We report three main results. First, PD patients with depression had globally attenuated alpha (8–13 Hz) rhythms, as well as attenuated central beta (13–30 Hz) and gamma (30–80 Hz) rhythms relative to PD patients without depression. Second, PD patients with depression had strong global differences in gamma rhythms relative to non-PD patients with depression. Third, we used a linear predictive coding of EEG Algorithm for PD (LEAPD) formulated by Anjum et al. [17,20], which provides binary classification based on resting-state EEG power spectra. LEAPD-based classification accurately identified PD patients with depression relative to PD patients and non-PD depressed patients. Collectively, these data implicate cortical rhythms in PD-related depression, which could lead to novel targeted therapies or new diagnostic biomarkers for this important non-motor aspect of PD.

## METHODS

### Participants

36 PD patients (11 women; Table S1) were recruited from clinics at the University of Iowa. A movement-disorders physician examined all PD patients to verify that they met the diagnostic criteria recommended by the United Kingdom PD Society Brain Bank criteria. Depression was quantified using the Geriatric Depression Scale in PD patients; a score of 5 to 15 was considered depressed). In addition, the motor Unified Parkinson’s Disease Rating Scale (UPDRS) was administered to all PD patients by a qualified rater, along with other clinical metrics, such as the Montreal Cognitive Assessment (MOCA) and behavioral assays. Data were collected with patients taking all medications as prescribed and PD patients were in the “ON” state. See our prior work for details of cognitive assessments [18]. Demographics and other clinical details are presented in Table S1 and were compared between groups by non-parametric Wilcoxon tests.

We recruited 12 demographically-similar depressed patients without PD (5 women; Table S1) from the University of Iowa’s depression and neuromodulation clinic. These patients were diagnosed with depression by the Patient Health Questionnaire-9, with a value of 9 to 27. A psychiatrist evaluated all patients, and patients took their medications as prescribed.

We obtained written informed consent from all participants according to the University of Iowa’s Institutional Review Board (IRB). Demographics of patients and control subjects are summarized in Table S1.

### EEG recording and analysis

Resting-state EEG was collected from patients while they sat in a quiet room with their eyes open for two minutes. Scalp EEG signals were collected from 64 channels of an EEG actiCAP (Brain Products GmbH) using a high-pass filter with a 0.1-Hz cutoff and a sampling frequency of 500 Hz. Electrode Pz was used as a reference, and electrode FPz was used as the ground. We used recording methods described previously in detail using a custom EEG cap with Iz, I1, and I2 leads in place of FT9, PO3, and PO4 leads; these leads were not analyzed [17–19]. We also removed FP1, FP2, FT10, TP9, and TP10 channels as these channels are often contamined by artifact, resulting in 56 channels for pre- and post-processing. EEG activity at the reference electrode Pz was recovered by computing the average reference. Bad channels and bad epochs were identified using the FASTER algorithm and the *pop_rejchan* function from EEGLAB and were then interpolated and rejected, respectively. Eye blinks were removed using independent component analysis (ICA). All channels were low-pass filtered at 100 Hz. Power was calculated using the *pwelch* function and was normalized to the mean power between 0–100 Hz for each channel. Scalp topography was plotted using *topoplot* from EEGLAB in delta (1-4 Hz), theta (4-8 Hz), alpha (8-13 Hz), beta (13-30 Hz), and gamma (30-80 Hz; Figure 1) bands.

**Figure 1:**
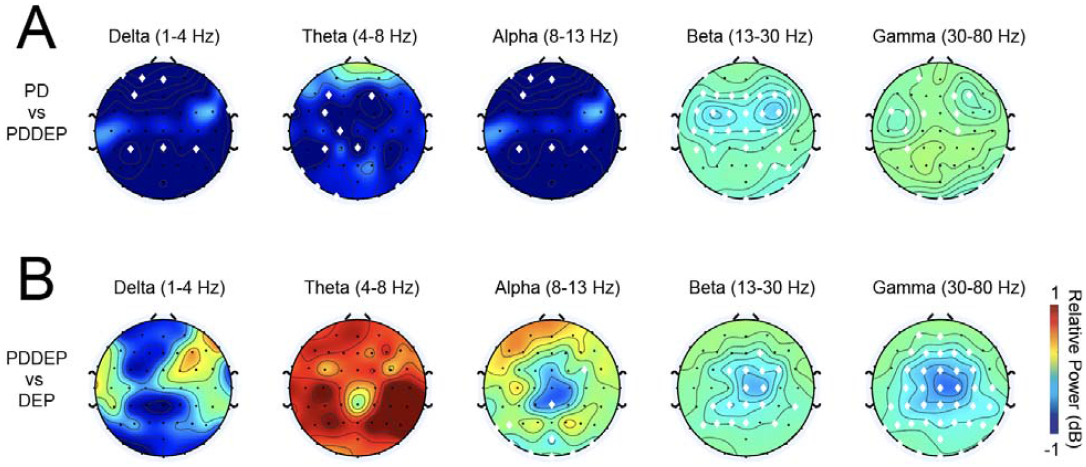
Scalp topography of relative EEG power in PD patients with depression. A) Relative power in PD patients with depression (PDDEP) compared to PD patients without depression for delta (1-4 Hz), theta (4-8 Hz), alph (8-13 Hz), beta (13-30 Hz), and gamma (30-80 Hz). B) Relative power in PDDEP compared to non-PD patients with depression (DEP). Electrodes are indicated by black dots; electrodes with significant differences between groups via ranksum testing are shown with white diamonds. Data from 18 PD, 18 PDDEP and 12 DEP.

### Machine learning using Linear predictive coding algorithms for PD (LEAPD)

LEAPD is an algorithm for binary classification of the spectral content of EEG signals. This approach was developed by Anjum et al. [17,20] to distinguish between PD patients and control participants. We implemented LEAPD to compare PD patients with depression (PDDEP) vs PD patients without depression (PD) and PDDEP vs depressed patients without PD (DEP). In particular, a LEAPD index between 0 and 1 is generated for each EEG recording, using the procedure outlined below. In each of the two problems, a threshold of 0.5 is used to distinguish between two groups. For example, if the LEAPD index for an EEG recording is below 0.5 then it is deemed to be in Group A and if above 0.5 it is classified as belonging to Group B.

In LEAPD, an EEG time series from a channel is processed using linear predictive coding (LPC) to encode the signal into coefficients of an autoregressive model minimizing the square of the prediction error [21] for that time series. The number of coefficients n, is called the LPC order. These coefficients are put in a vector of dimension n with one entry for each coefficient. An LPC vector is generated by substracting the mean. Each LPC vector is viewed as a point in the n-dimensional space. LPC vectors of each group lies on distinct affine subspaces. For example those for PDDEP roughly lie on one affine subspace while those of PD on another. An affine subspace is the generalization of a one-dimensional line or a two-dimensional plane in larger dimensions. The LEAPD index of a recording is as below, where Dl is the distance of its LPC vector from the affine subspace of one group and D2 is the distance from the affine subspace of the other group:

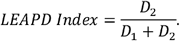

Principal Component Analysis (PCA) is used to identify the affine subspace of a given dimension that best fits the LPC vectors of each group. Parameters used to control the learning process include: (1) the cutoff frequencies of the filter used to process the EEG data; (2) the length of the LPC vector (LPC order); and (3) the dimension of the affine subspace.

We quantified differences between LEAPD values for each channel using non-parametric Wilcoxon *ranksum* tests. In addition, we used a classifier to calculate the accuracy of PD vs PDDEP and DEP vs PD at each channel. Two-channel LEAPD values were computed by taking the geometric mean of the LEAPD values for each channel. We then used a classifier on all two-channel combinations, and we presented results only from selected high-performing combinations.

As the dataset was small, we could not perform out-of-sample prospective tests to validate the accuracy of the model. However, we tested the robustness of the results by examining LEAPD performance on truncated data. In all instances leave-one-out cross validation (LOOCV) was used to quantify performance. LOOCV uses the entire dataset without one test sample to predict each test sample, which protects against the overfitting common with small datasets. We report data from individual channels and combinations of channels that yielded the a) highest accuracy in discriminating PD vs PDDEP and PDDEP vs DEP, and b) were the most robust on truncated data.

## RESULTS

PD patients with and without depression had similar age (*p* = 0.23), motor function as measured by UPDRS (*p* = 0.22), and cognitive profiles as measured by the MOCA (*p* = 0.94 value; Table S1). We collected resting-state EEG data and compared scalp topography of relative power for PD patients vs PD patients with depression (PDDEP) at delta (1-4 Hz), theta (4-8 Hz), alpha (8–13 Hz), beta (13–30 Hz), and gamma bands (30–80 Hz; Figure 1A). We also compared scalp topography for PDDEP vs non-PD patients with depression (DEP; Figure 1B). These data illustrate that there can be band-specific differences that distinguish depression in PD.

Our machine learning approach, LEAPD, compress power spectra into a series of autoregressive coefficients that holistically captures the shape of each power spectra with a few numbers [17,20]. Here, we used LEAPD to classify PD vs PDDEP and PDDEP vs DEP from single channels, as well as combinations of two channels (Figure 2).

**Figure 2:**
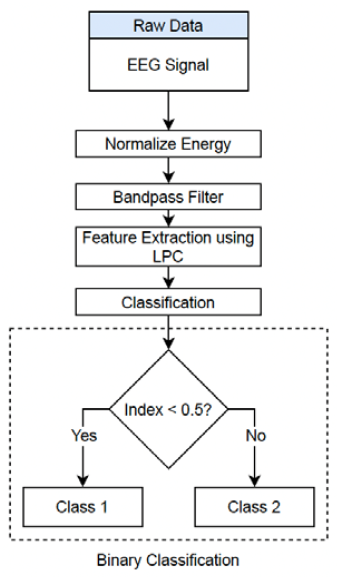
LEAPD Classification approach: Flow chart of classification.

We first used LEAPD to discriminate 18 PD from 18 PDDEP patients across all EEG electrodes (Figure 3A). Single-channel accuracy for channel CP3 was 86% and for TP8 was 86% (Figure 3A). Combining both CP3 and TP8 resulted in an overall LOOCV classification accuracy of 97%. These channels had distinct LEAPD indices between PD and PDDEP (CP3: *p* = 0. 00009, Cohen’s d = 1.8; TP8: *p* = 0. 0.00004, Cohen’s d = 1.8; CP3+TP8: *p* < 0.001; Cohen’s d = 3.25; Figure 3B). Receiver-operator curves (ROCs) for these channels in predicting PD vs PDDEP are shown in Figure 3C.

**Figure 3:**
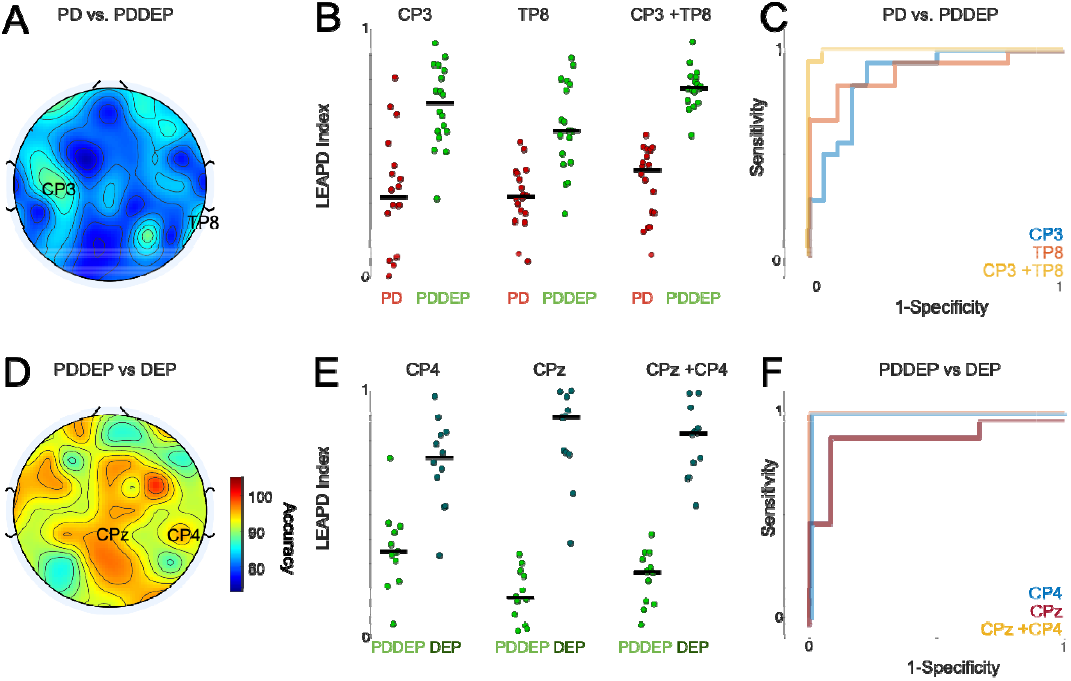
Machine-learning classification of LEAPD. A) We constructed LEAPD indices from LPC coefficients from electrodes CP3 and TP8 for PD patients without depression (PD) vs PD patients with depression (PDDEP). B) Receiv r-operating curves (ROC) for single-channel performance of CP3, TP8, and CP3+TP8 combined, and C) channel performance across single electrodes. Data from 18 PD and 18 PDDEP patients. D) We also generated LEAPD indices for PDDEP (green) compared to depressed patients without PD (DEP; dark green) at CP4, CPz, and CP4+CPz combined. E) ROC curves and F) single channel performance across single electrodes. Data from 12 PDDEP and 12 DEP patients.

In addition, we found that LEAPD was highly accurate in differentiating 12 PDDEP patients (selected at random from 18 total) from 12 DEP patients, with 96% single-channel signal accuracy for electrode CPz and 92% for electrode CP4. Combining both channels resulted in 100% classification accuracy (Figure 3F). For these electodes, LEAPD distguished PDDEP vs DEP (Figure 3D; CPz: *p* = 0.00004, Cohen’s d: = 4.3; CP4: *p* = 0.0007, Cohen’s d: 2.0; CP4+CPz: *p* = 0.0004; Cohen’s d = 4.3; Figure 3E).

Additionally, we performed a truncation analysis of CP3, TP8, and CP3+TP8 combined for PD vs PDDEP and of CPz, CP4, and CPz+CP4 combined for DEP vs PDDEP. Recorded EEG data were truncated from full-length samples to samples that were a fraction of the original length. LEAPD analysis was then performed on the shortened signal using the same hyperparameters as those of the original signal. Truncation fractions of 0.05, 0.33 and 0.67 were tested. Performance of the channels at each truncation fraction is shown in Table S2. Although truncation did reduce the accuracy of the channels, each channel still retained significant discriminatory ability at shorter signal lengths. The performance degraded gracefully with truncation, indicating that the signals chosen are likely measuring a fundamental difference in EEG behavior between classes, rather than an artifact of overfitting. It is notable that accuracy of greater than 85% was achieved from two minutes of resting-state EEG signals. Performance on truncated data is shown in Figure 4 for channels of interest for PD vs PDDEP (Figure 4A) and PDDEP vs DEP (Figure 4B). Collectively, these data suggest that spectral features of scalp EEG can distinguish depression in PD.

**Figure 4:**
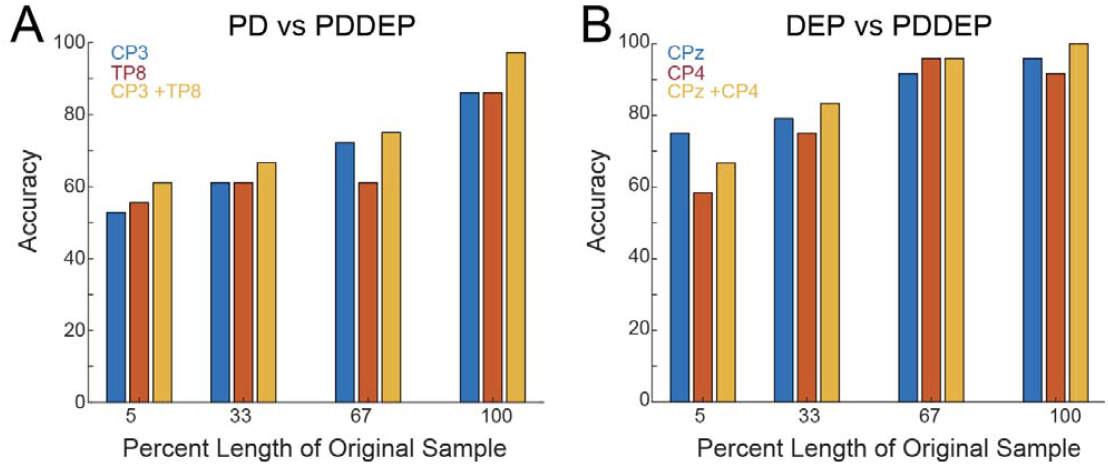
Truncation analysis of LEAPD-based classification. A) Data from PD vs PDDEP for 5, 33, 67, and 100% of data for PDDEP vs PD for CP3, TP8, and CP3+TP8 and B) PDDEP vs DEP for CP4, CPz, and CP4 + CPz.

## DISCUSSION

We explored the cortical basis of depression in PD using resting-state scalp EEG. We found that PD patients with depression had central differences in beta and gamma rhythms. We used LEAPD, a spectral machine-learning approach, to detect differences in EEG signals from two minutes of resting-state data from a single electrode, achieving accuracies of 97% for PD patients with and without depression and 100% for PD vs non-PD patients with depression.These data indicate that PD patients with depression can be accurately differentiated from PD patients without depression and from depressed non-PD patients using machine learning.

Depression is a complex disorder [22] involving many brain networks; however, one consistent finding is abnormal cortical function [14,23]. Scalp EEG studies have found dysfunctional alpha rhythms in depressed patients [24,25], a finding that we report here comparing PD patients with and without depression. Beta rhythms can be profoundly abnormal in PD [26] and our data here indicate that depression decreases resting-state beta, alpha, and gamma rhythms in PD. We find that many cortical regions are implicated in PD-related depression, including prefrontal and parietal regions that have been found in prior studies of depression [14,27]

These data suggest that EEG, which is relatively inexpensive and ubiquitously available, can identify PD patients with depression. This is important because depression can be missed in PD [3–5], and electrophysiological diagnostic tools may aid in this effort. We report that our spectral approach can rapidly, robustly, and accurately identify EEG signals from PD patients with depression. Our results are in line with previous efforts to use LEAPD to identify local field potentials from animal models of PD and EEG data recorded from PD patients and controls [17,20]. LEAPD-based techniques might have additional utility in settings where neurophysiology is common, such as during deep-brain stimulation surgeries, and they may be helpful for closed-loop control applications. Apart from being robust and accurate, LEAPD is amenable to fast implementation and can serve as a trigger mechanism for brain stimulation.

Our work is supported by prior qEEG studies describing that a single parameter can differentiate depression and dementia in PD [15]. An early study which averages across all EEG electrodes reported distinct scalp topography of depressed PD patients, focusing on alpha rhythms [7]. Our study is supportive of these differences, and we are able to localize these results to the left frontal electrodes. In addition, we find broader differences over central electrodes in beta and gamma bands, which may have been averaged out in prior work that averaged EEG signals from multiple electrodes. Finally, we used advanced machine-learning to distinguish PD patients with depression from both PD patients and non-PD patients with depression. Recent work has reported frontal differences in sleep in PD patients with depression[28], as well as differences between midline event-related potentials between PD patients with and without depression [29]. Our study extends these findings and helps define the spectral topography of resting-state EEG in PD patients with depression, and demonstrates the potential of machine-learning for identifying PD patients with depression.

In this manuscript, we illustrate these effects from relatively high-performing channels: CP3/TP8 in PD vs PDDEP and CP4/CPz in PDDEP vs DEP. We chose these exemplars to illustrate high-performing channel combinations from each comparison. However, we note that channels also had high performance, and could be used for classification and identificiation of depression in PD.

Our study has several limitations. First, our sample size was limited, although in line with prior EEG studies in PD patients with depression [28,29]. Second, all of our patients were medicated, and it is possible that medications could influence these EEG signals [30]. Third, our method of diagnosing depression and quantifying symptom burden in PD patients was distinct from the method used with non-PD patients, limiting comparisons between these groups. Finally, our LEAPD approach did not include an out-of-sample prospective test, though the truncation analysis does remove concerns of overfitting. Despite these shortfalls, our findings describe spectral changes in PD patients with depression compared to PD patients without depression and non-PD patients with depression. We report that LEAPD-based machine learning approaches can identify EEG signals from PD patients with depression. These data could help illuminate the cortical neurophysiology of PD-related depression and could help lead to new biomarkers or diagnostic tools.

## Data Availability

All data produced in the present study are available upon reasonable request to the authors

## AUTHOR DECLARATION

None of the authors have any potential conflicts of interest to disclose.

## ACKNOWLEDGEMENTS

This data was supported by NIH P20NS123151 and R01NS100849 to NSN.

## Supplementary Tables

**Table S1:**
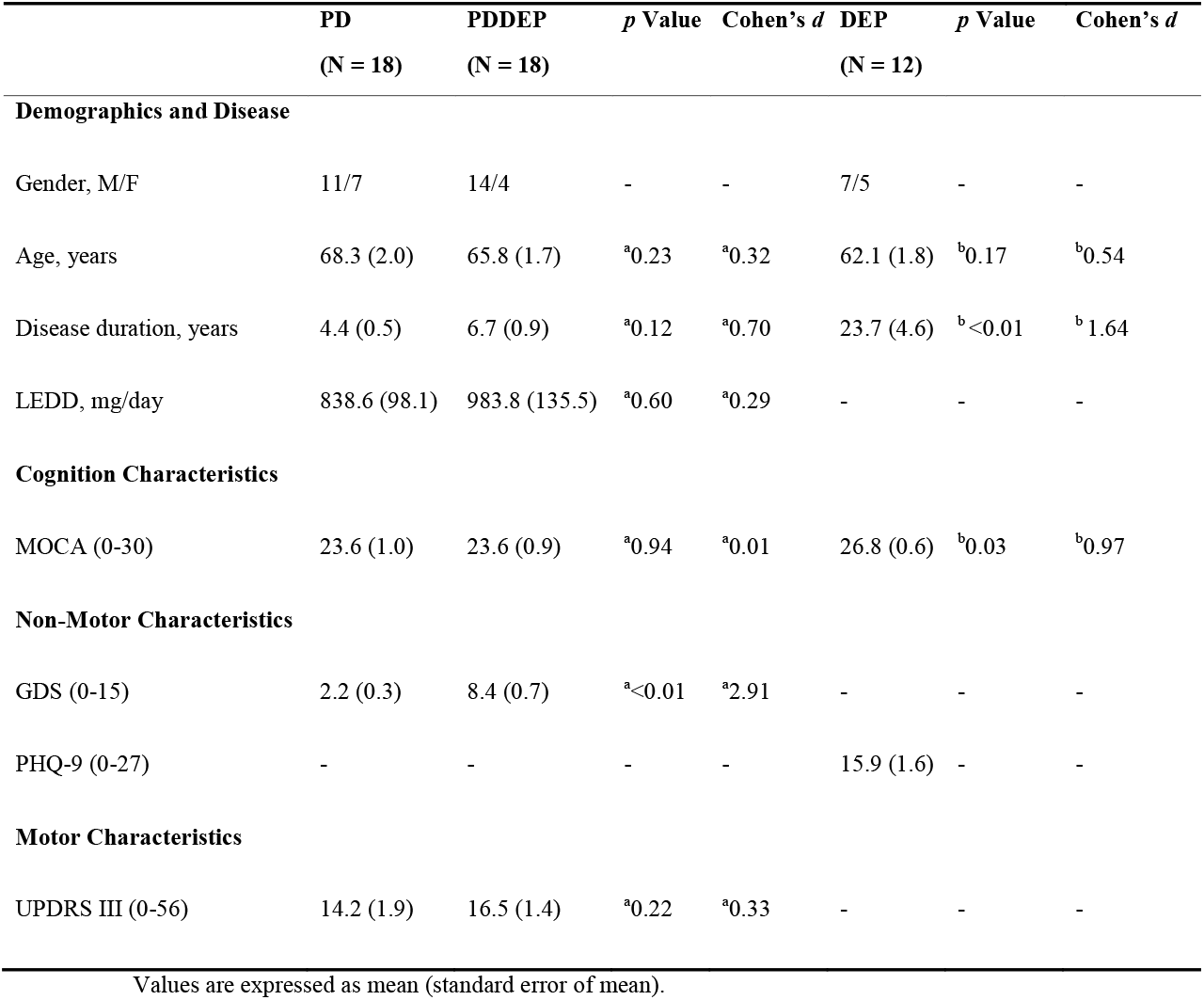
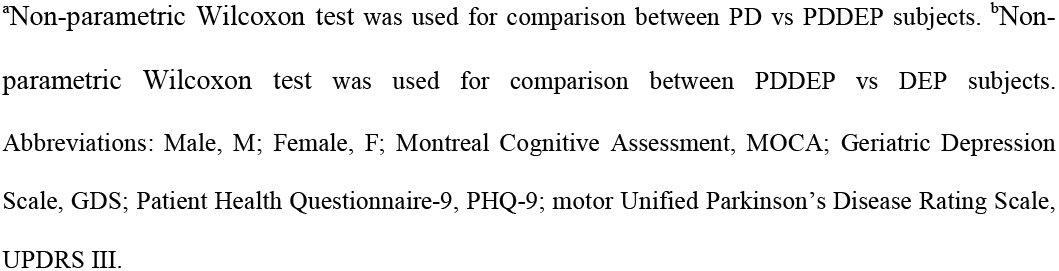
Demographic, disease, non-motor, motor, and cognitive characteristics.

|  | **PD**<br>(N = 18) | **PDDEP**<br>(N = 18) | *p* Value | Cohen's *d* | **DEP**<br>(N = 12) | *p* Value | Cohen's *d* |
| --- | --- | --- | --- | --- | --- | --- | --- |
| **Demographics and Disease** |  |  |  |  |  |  |  |
| Gender, M/F | 11/7 | 14/4 | - | - | 7/5 | - | - |
| Age, years | 68.3 (2.0) | 65.8 (1.7) | <sup>a</sup>0.23 | <sup>a</sup>0.32 | 62.1 (1.8) | <sup>b</sup>0.17 | <sup>b</sup>0.54 |
| Disease duration, years | 4.4 (0.5) | 6.7 (0.9) | <sup>a</sup>0.12 | <sup>a</sup>0.70 | 23.7 (4.6) | <sup>b</sup><0.01 | <sup>b</sup>1.64 |
| LEDD, mg/day | 838.6 (98.1) | 983.8 (135.5) | <sup>a</sup>0.60 | <sup>a</sup>0.29 | - | - | - |
| **Cognition Characteristics** |  |  |  |  |  |  |  |
| MOCA (0-30) | 23.6 (1.0) | 23.6 (0.9) | <sup>a</sup>0.94 | <sup>a</sup>0.01 | 26.8 (0.6) | <sup>b</sup>0.03 | <sup>b</sup>0.97 |
| **Non-Motor Characteristics** |  |  |  |  |  |  |  |
| GDS (0-15) | 2.2 (0.3) | 8.4 (0.7) | <sup>a</sup><0.01 | <sup>a</sup>2.91 | - | - | - |
| PHQ-9 (0-27) | - | - | - | - | 15.9 (1.6) | - | - |
| **Motor Characteristics** |  |  |  |  |  |  |  |
| UPDRS III (0-56) | 14.2 (1.9) | 16.5 (1.4) | <sup>a</sup>0.22 | <sup>a</sup>0.33 | - | - | - |

Values are expressed as mean (standard error of mean).
<sup>a</sup>Non-parametric Wilcoxon test was used for comparison between PD vs PDDEP subjects. <sup>b</sup>Non-parametric Wilcoxon test was used for comparison between PDDEP vs DEP subjects. Abbreviations: Male, M; Female, F; Montreal Cognitive Assessment, MOCA; Geriatric Depression Scale, GDS; Patient Health Questionnaire-9, PHQ-9; motor Unified Parkinson's Disease Rating Scale, UPDRS III.

**Table S2:**
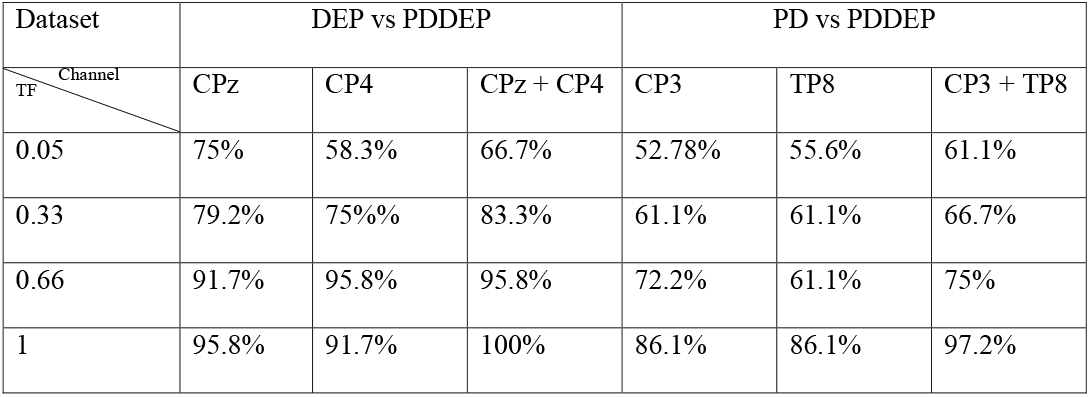
Truncation analysis accuracy across datasets.

| Dataset | DEP vs PDDEP |  |  | PD vs PDDEP |  |  |
| --- | --- | --- | --- | --- | --- | --- |
| Channel<br>TF | CPz | CP4 | CPz + CP4 | CP3 | TP8 | CP3 + TP8 |
| 0.05 | 75% | 58.3% | 66.7% | 52.78% | 55.6% | 61.1% |
| 0.33 | 79.2% | 75%% | 83.3% | 61.1% | 61.1% | 66.7% |
| 0.66 | 91.7% | 95.8% | 95.8% | 72.2% | 61.1% | 75% |
| 1 | 95.8% | 91.7% | 100% | 86.1% | 86.1% | 97.2% |

## References

[1] K.R. Chaudhuri, P. Odin, The challenge of non-motor symptoms in Parkinson’s disease, Prog. Brain Res. 184 (2010) 325–341. https://doi.org/10.1016/S0079-6123(10)84017-8.

[2] A. Lieberman, Depression in Parkinson’s disease – a review, Acta Neurol. Scand. 113 (2006) 1–8. https://doi.org/10.1111/j.1600-0404.2006.00536.x.

[3] H. Allain, S. Schuck, N. Maudui, Depression in Parkinson’s disease: Must be properly diagnosed and treated to avoid serious morbidity, BMJ. 320 (2000) 1287–1288. https://doi.org/10.1136/bmj.320.7245.1287.

[4] S. Muzerengi, H. Lewis, M. Edwards, E. Kipps, A. Bahl, P. Martinez-Martin, K.R. Chaudhuri, Non-motor symptoms in Parkinson’s disease: an underdiagnosed problem, Aging Health. 2 (2006) 967–982. https://doi.org/10.2217/1745509X.2.6.967.

[5] M.H.M. Timmer, M.H.C.T. van Beek, B.R. Bloem, R.A.J. Esselink, What a neurologist should know about depression in Parkinson’s disease, Pract. Neurol. 17 (2017) 359–368. https://doi.org/10.1136/practneurol-2017-001650.

[6] A.F.G. Leentjens, M. Van den Akker, J.F.M. Metsemakers, R. Lousberg, F.R.J. Verhey, Higher incidence of depression preceding the onset of Parkinson’s disease: a register study, Mov. Disord. Off. J. Mov. Disord. Soc. 18 (2003) 414–418. https://doi.org/10.1002/mds.10387.

[7] S.R. Filipović, N. Čovičković-Šternić, M. Stojanović-Svetel, D. Lečić, V.S. Kostić, Depression in Parkinson’s disease: an EEG frequency analysis study, Parkinsonism Relat. Disord. 4 (1998) 171–178. https://doi.org/10.1016/S1353-8020(98)00027-3.

[8] D. Aarsland, S. Påhlhagen, C.G. Ballard, U. Ehrt, P. Svenningsson, Depression in Parkinson disease—epidemiology, mechanisms and management, Nat. Rev. Neurol. 8 (2012) 35–47. https://doi.org/10.1038/nrneurol.2011.189.

[9] N.L. Bormann, N.T. Trapp, N.S. Narayanan, A.D. Boes, Developing Precision Invasive Neuromodulation for Psychiatry, J. Neuropsychiatry Clin. Neurosci. 33 (2021) 201–209. https://doi.org/10.1176/appi.neuropsych.20100268.

[10] N.S. Narayanan, R.L. Rodnitzky, E.Y. Uc, Prefrontal dopamine signaling and cognitive symptoms of Parkinson’s disease, Rev. Neurosci. 24 (2013) 267–278. https://doi.org/10.1515/revneuro-2013-0004.

[11] S. Ghosal, B.D. Hare, R.S. Duman, Prefrontal cortex GABAergic deficits and circuit dysfunction in the pathophysiology and treatment of chronic stress and depression, Curr. Opin. Behav. Sci. 14 (2017) 1–8. https://doi.org/10.1016/j.cobeha.2016.09.012.

[12] Y.-C. Kim, N.S. Narayanan, Prefrontal D1 Dopamine-Receptor Neurons and Delta Resonance in Interval Timing., Cereb. Cortex N. Y. N 1991. (2018). https://doi.org/10.1093/cercor/bhy083.

[13] A.Y. Deutch, Prefrontal cortical dopamine systems and the elaboration of functional corticostriatal circuits: implications for schizophrenia and Parkinson’s disease, J. Neural Transm. Gen. Sect. 91 (1993) 197–221.

[14] M.S. George, T.A. Ketter, R.M. Post, Prefrontal cortex dysfunction in clinical depression, Depression. 2 (1994) 59–72. https://doi.org/10.1002/depr.3050020202.

[15] A. Primavera, P. Novello, Quantitative electroencephalography in Parkinson’s disease, dementia, depression and normal aging, Neuropsychobiology. 25 (1992) 102–105. https://doi.org/10.1159/000118817.

[16] M.S. George, Z. Nahas, M. Molloy, A.M. Speer, N.C. Oliver, X.-B. Li, G.W. Arana, S.C. Risch, J.C. Ballenger, A controlled trial of daily left prefrontal cortex TMS for treating depression, Biol. Psychiatry. 48 (2000) 962–970. https://doi.org/10.1016/S0006-3223(00)01048-9.

[17] M.F. Anjum, S. Dasgupta, R. Mudumbai, A. Singh, J.F. Cavanagh, N.S. Narayanan, Linear predictive coding distinguishes spectral EEG features of Parkinson’s disease, Parkinsonism Relat. Disord. 79 (2020) 79–85. https://doi.org/10.1016/j.parkreldis.2020.08.001.

[18] A. Singh, R.C. Cole, A.I. Espinoza, A. Evans, S. Cao, J.F. Cavanagh, N.S. Narayanan, Timing variability and midfrontal ∼4 Hz rhythms correlate with cognition in Parkinson’s disease, NPJ Park. Dis. 7 (2021) 14. https://doi.org/10.1038/s41531-021-00158-x.

[19] A. Singh, R.C. Cole, A.I. Espinoza, D. Brown, J.F. Cavanagh, N.S. Narayanan, Frontal theta and beta oscillations during lower-limb movement in Parkinson’s disease, Clin. Neurophysiol. Off. J. Int. Fed. Clin. Neurophysiol. 131 (2020) 694–702. https://doi.org/10.1016/j.clinph.2019.12.399.

[20] M.F. Anjum, J. Haug, S.L. Alberico, S. Dasgupta, R. Mudumbai, M.A. Kennedy, N.S. Narayanan, Linear Predictive Approaches Separate Field Potentials in Animal Model of Parkinson’s Disease, Front. Neurosci. 14 (2020) 394. https://doi.org/10.3389/fnins.2020.00394.

[21] B.S. Atal, The history of linear prediction, IEEE Signal Process. Mag. 23 (2006) 154–161. https://doi.org/10.1109/MSP.2006.1598091.

[22] M. aan het Rot, S.J. Mathew, D.S. Charney, Neurobiological mechanisms in major depressive disorder, CMAJ Can. Med. Assoc. J. 180 (2009) 305–313. https://doi.org/10.1503/cmaj.080697.

[23] S. Zhao, J. Kong, S. Li, Z. Tong, C. Yang, H. Zhong, Randomized controlled trial of four protocols of repetitive transcranial magnetic stimulation for treating the negative symptoms of schizophrenia, Shanghai Arch. Psychiatry. 26 (2014) 15–21. https://doi.org/10.3969/j.issn.1002-0829.2014.01.003.

[24] I.H. Gotlib, EEG Alpha Asymmetry, Depression, and Cognitive Functioning, Cogn. Emot. 12 (1998) 449–478. https://doi.org/10.1080/026999398379673.

[25] R. Thibodeau, R.S. Jorgensen, S. Kim, Depression, anxiety, and resting frontal EEG asymmetry: A meta-analytic review, J. Abnorm. Psychol. 115 (2006) 715–729. https://doi.org/10.1037/0021-843X.115.4.715.

[26] N. Jenkinson, P. Brown, New insights into the relationship between dopamine, beta oscillations and motor function, Trends Neurosci. 34 (2011) 611–618. https://doi.org/10.1016/j.tins.2011.09.003.

[27] J.L. Stewart, D.N. Towers, J.A. Coan, J.J.B. Allen, The oft-neglected role of parietal EEG asymmetry and risk for major depressive disorder, Psychophysiology. 48 (2011) 82–95. https://doi.org/10.1111/j.1469-8986.2010.01035.x.

[28] K. Liu, Q. Ma, M. Wang, Comparison of Quantitative Electroencephalogram During Sleep in Depressed and Non-Depressed Patients with Parkinson’s Disease, Med. Sci. Monit. Int. Med. J. Exp. Clin. Res. 25 (2019) 1046–1052. https://doi.org/10.12659/MSM.913931.

[29] N.N.W. Dissanayaka, T.R. Au, A.J. Angwin, K.K. Iyer, J.D. O’Sullivan, G.J. Byrne, P.A. Silburn, R. Marsh, G.D. Mellick, D.A. Copland, Depression symptomatology correlates with event-related potentials in Parkinson’s disease: An affective priming study, J. Affect. Disord. 245 (2019) 897–904. https://doi.org/10.1016/j.jad.2018.11.094.

[30] R. Aiyer, V. Novakovic, R.L. Barkin, A systematic review on the impact of psychotropic drugs on electroencephalogram waveforms in psychiatry, Postgrad. Med. 128 (2016) 656–664. https://doi.org/10.1080/00325481.2016.1218261.

